# Students’ Perceptions of an AI-Enhanced Ethics Learning Platform: A Pilot Study on Interprofessional Healthcare Education

**DOI:** 10.64898/2026.06.23.26356394

**Authors:** Liam Rankine, Jennifer Van Bussel, Sheila Theresa Moodie, Andrews K Tawiah

## Abstract

**Introduction:** Generative artificial intelligence (AI) can produce realistic clinical scenarios on demand and deliver immediate, individualized feedback, yet its use to teach ethical reasoning, rather than to address the ethics of AI itself, remains underexplored in interprofessional healthcare education.

**Aim:** This pilot study examined how interprofessional healthcare students perceived an AI-enhanced, case-based platform designed to support ethical decision-making across physical therapy, occupational therapy, speech-language pathology, and audiology.

**Methods:** Students enrolled in an interprofessional education course completed an online module of 20 instructor-vetted, AI-generated ethics cases and an optional post-activity survey of Likert-scale and open-ended items. Quantitative data were analyzed descriptively and qualitative responses were analyzed through content analysis.

**Results:** Ten students responded. Within this small sample, perceptions of platform utility and usability were strongly positive, with all respondents agreeing that immediate feedback and scenario variety supported learning. Perceptions were more divided when the platform was compared directly with traditional classroom learning, and respondents identified pacing and auto-scrolling as usability concerns.

**Conclusions:** These preliminary findings suggest AI-enhanced case-based platforms can engage students and support applied ethics learning but are best positioned to complement rather than replace traditional instruction. Findings are exploratory given the small, demographically limited sample.

## 1. Introduction

Artificial intelligence (AI), and generative AI (GenAI) in particular, has rapidly transformed higher education by enabling dynamic, adaptive learning environments that extend beyond static instructional materials [1,2]. Large language models (LLMs) can generate diverse, realistic scenarios on demand, provide immediate personalized feedback, and support self-paced, interactive engagement as students develop professional competencies across disciplines.

Ethics education within interprofessional healthcare programs presents a compelling use case for AI-enhanced approaches. Students in physical therapy, occupational therapy, speech-language pathology, and audiology must be prepared to navigate nuanced ethical situations involving patient care, professional responsibility, and interprofessional collaboration. Traditional classroom approaches, which typically rely on static case studies and group discussion, are limited in their ability to expose students to a broad range of scenarios, provide timely individualized feedback, or replicate the complexity of real-world ethical dilemmas [3,4]. Time and curriculum constraints further restrict the depth of engagement possible within formal class time [5,6].

A growing body of evidence supports integrating GenAI into case-based and problem-based learning. A systematic review and meta-analysis of AI-powered case-based learning in medical and dental education reported a 46% improvement in knowledge acquisition [7]. AI-supported case discussions have been associated with significant gains in student self-efficacy, attributed to adaptive feedback, dynamic modelling, and increased engagement [8]. AI can also efficiently generate realistic clinical case materials, though human review remains essential to ensure accuracy and clinical relevance [9], and AI-generated interactive personas can transform static cases into immersive, inquiry-driven experiences that deepen strategic thinking and stakeholder analysis [10]. However, deploying such tools without adequate oversight can perpetuate algorithmic bias, widen the digital divide, and compromise data privacy [11,12].

Despite this growing evidence base, the literature remains concentrated within technical, medical, and skills-based domains. Much of the discussion of AI and ethics focuses on the ethical issues of using GenAI (academic integrity, bias, and data governance) rather than on using AI as a means to teach ethical reasoning. There is limited exploration of how GenAI can be intentionally designed to support ethical decision-making through case-based learning. Ethical decision-making poses unique challenges relative to factual or procedural knowledge, requiring critical thinking, perspective-taking, and evaluation of ambiguous and value-laden scenarios. These characteristics are well aligned with the interactive, adaptive capabilities of GenAI, yet this application remains underexplored. This paper addresses that gap by reporting preliminary pilot data on student perceptions of an AI-enhanced, case-based learning platform designed to support ethical reasoning in an interprofessional healthcare education setting; all AI-generated scenarios were reviewed and vetted by course instructors prior to deployment.

## 2. Aims

This study explored how interprofessional healthcare students (physical therapy, occupational therapy, speech-language pathology, and audiology) perceive an AI-enhanced interactive learning platform for developing knowledge of ethical decision-making. Three objectives guided the work: (1) to examine whether the AI-generated platform benefits student understanding and learning of ethical concepts; (2) to identify which platform features (e.g., case realism, immediate feedback, pacing) students find most valuable or challenging for learning; and (3) to compare students’ perceptions of AI-driven case discussions with their experiences of traditional case-based learning in their programs.

## 3. Methods

### Study design and setting

This pilot study used a cross-sectional survey design. Interprofessional healthcare students enrolled in a required interprofessional education course completed an online AI-enhanced ethics learning module and were then invited to complete a voluntary post-activity survey comprising Likert-scale items (technology acceptance, engagement, convenience, and perceived learning support) and several open-ended questions eliciting reflections on the platform’s strengths, weaknesses, and suggested improvements.

### AI-enhanced learning platform

As illustrated in Figure 1, the development pipeline followed an iterative process of AI-assisted generation and instructor review. The platform comprised 20 clinical case scenarios across four professions: physical therapy, occupational therapy, speech-language pathology, and audiology. Cases were presented through a chat-based interface with animated dialogue bubbles and emoji avatars representing healthcare professionals. For each scenario, students identified whether the case represented **ethical uncertainty**, an **ethical dilemma**, or **ethical distress**; the platform then provided immediate, tailored feedback explaining the correct answer and a reflection prompt encouraging deeper engagement. Students typically completed the module in approximately 8–10 minutes. The platform was embedded within the institutional learning management system. All case scenarios were generated using a large language model and were subsequently reviewed and approved by the course instructors prior to deployment. The platform source code is publicly available [URL withheld for peer review].

**Figure 1.**
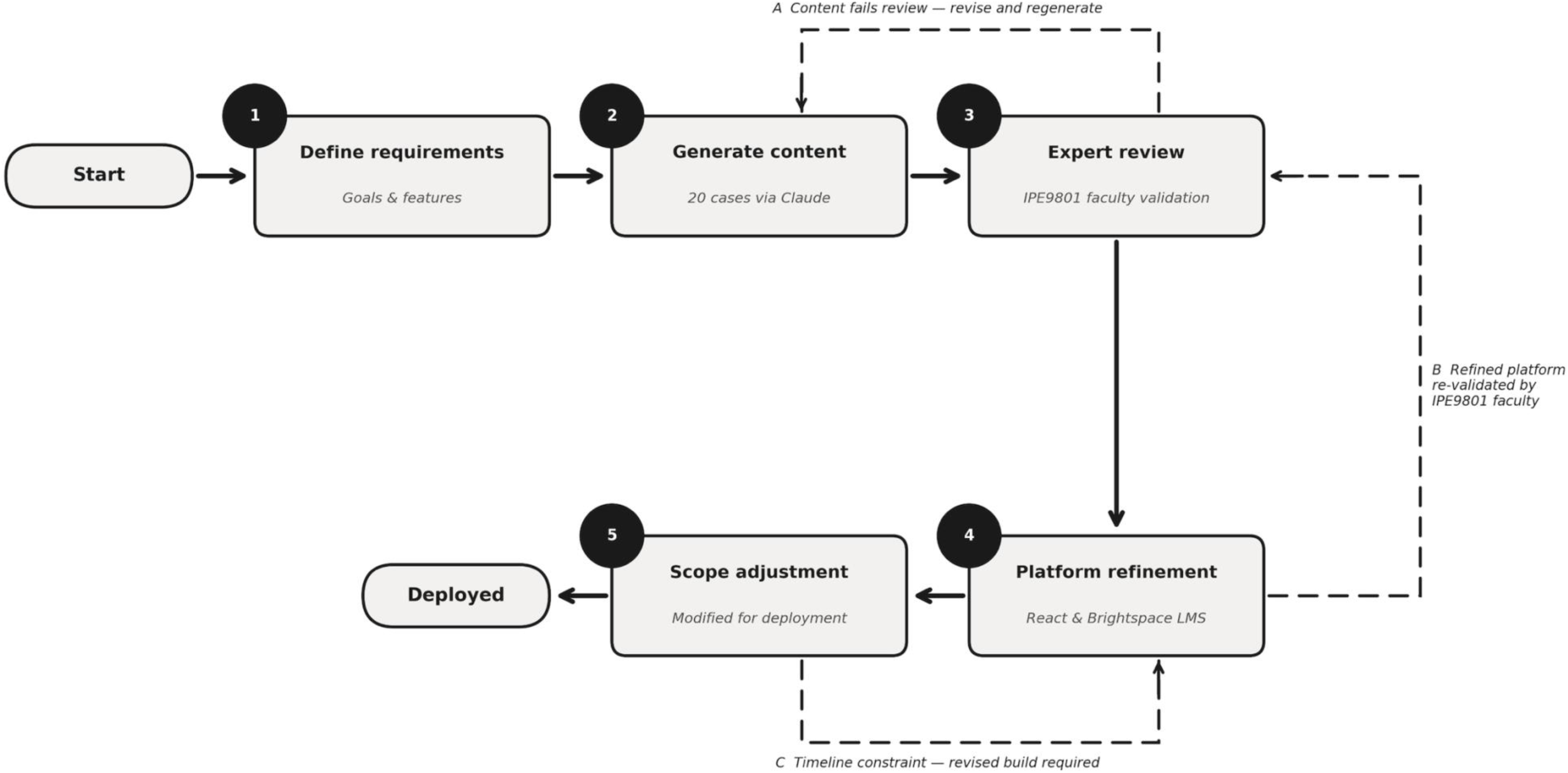
Development pipeline for the AI-enhanced ethics learning platform

### Questionnaire development and administration

Data were collected using a structured post-activity questionnaire developed in the institutional Qualtrics system. The instrument included demographic items (program, gender, race/ethnicity, and age) to characterize the sample, and Likert-scale items assessing perceived usefulness for learning, engagement, convenience, ease of use, and comparison with traditional learning generally and with other case-based learning. Additional items probed usability and user experience. Open-ended questions invited participants to describe what they liked most and least, how the platform influenced their ethical reasoning, and suggestions for improvement. The questionnaire was developed by the course instructor to ensure clarity, alignment with course objectives, and coverage of learning, convenience, and comparison with other learning paradigms.

### Population, sampling, and recruitment

The target population was interprofessional healthcare students enrolled in the course (approximately 318 students across the four programs). The intended sample was 175 students expected to complete the module and provide informed consent. Students were recruited through course announcements and email invitations distributed by the instructor. Recruitment materials included a study description, an information letter, and a link and Quick Response (QR) code to the platform presented at the end of the module. Participation was voluntary, had no impact on course grades or standing, and could be discontinued at any time before submission. Prior to data collection, the platform and questionnaire were pilot checked with instructor oversight to ensure technical reliability, alignment with course objectives, and clarity of survey items.

### Ethics

The study received approval from the institutional non-medical research ethics board, in accordance with institutional policies and the Tri-Council Policy Statement on research involving humans. Students reviewed an electronic Letter of Information and indicated consent before beginning the survey; no directly identifying information was collected.

### Data analysis

Quantitative survey data were exported to a spreadsheet for descriptive analysis, including means and frequency distributions for Likert-scale items relating to the three objectives. Given the small sample, exploratory comparisons across programs and demographic groups were not conducted. Open-ended responses were analyzed through content analysis, identifying recurring patterns in comments about perceived learning value, usability, and comparison with other learning paradigms.

## 4. Results

This pilot study targeted 175 respondents among the 318 enrolled students (∼55% expected response rate); however, only 10 students responded (∼3% actual response rate). The low response rate reflects survey distribution at the end of term, coinciding with the start of the examination period. The small sample substantially limits the statistical reliability and generalizability of the findings, which should be interpreted as preliminary and exploratory; nonetheless, participants provided qualitative data of value for future use of AI-enabled learning platforms.

### Sample characteristics

Table 1 presents respondent demographics. All 10 respondents identified as women, so the sample does not capture the perspectives of male or non-binary students. Most participants self-identified as White/Caucasian, with the remainder distributed roughly equally across African/Caribbean, East Asian, and Middle Eastern/West Asian backgrounds. Most respondents were enrolled in audiology, with the remainder in speech-language pathology and occupational therapy. Notably, no physiotherapy students completed the survey; the findings therefore reflect the perspectives of audiology, speech-language pathology, and occupational therapy students only and should not be interpreted as representative of all four target professions.

**Table 1.** Demographic characteristics of survey respondents (N = 10).

| Characteristic | Category | Frequency (n) | Percentage (%) |
| --- | --- | --- | --- |
| Gender | Female | 10 | 100 |
| Age (years) | 22 | 2 | 20 |
|  | 23 | 2 | 20 |
|  | 24 | 3 | 30 |
|  | 26 | 1 | 10 |
|  | 27 | 2 | 20 |
| Ethnicity | African/Caribbean | 1 | 10 |
|  | White/Caucasian | 5 | 50 |
|  | East Asian <sup>a</sup> | 3 | 30 |
|  | Middle Eastern/West Asian <sup>b</sup> | 1 | 10 |
| Area of study | Audiology | 5 | 50 |
|  | Speech-Language Pathology | 2 | 20 |
|  | Occupational Therapy | 3 | 30 |

<sup>a</sup> East Asian includes China, Taiwan, Hong Kong, Japan, and Korea. <sup>b</sup> Middle Eastern/West Asian includes Afghanistan, Iran, Saudi Arabia, and Syria.

### Objective 1: Platform utility and ethical concept learning

Six of the 16 survey items addressed Objective 1, evaluating whether the platform improved understanding of ethical practice concepts and strengthened students’ ability to differentiate ethical uncertainty, dilemmas, and distress (Figure 2). Within this small sample, perceptions were strongly positive: agreement across all six items ranged from 80% to 100%, with no neutral responses. Two items reached 100% agreement, that immediate feedback was useful for understanding ethical practice, and that scenario variety was valuable for learning. Two further items reached 90% agreement (relevance to real ethical practice; improved ability to reason through ethical problems). The two items concerning improvement in distinguishing the specific ethical concepts had the lowest agreement (80%). Qualitatively, participants noted that immediate feedback and scenario variety enhanced learning; one described new examples with timely responses that helped prepare for an examination, and another described the platform as engaging and detailed in helping differentiate the three types of ethical challenge.

**Figure 2.**
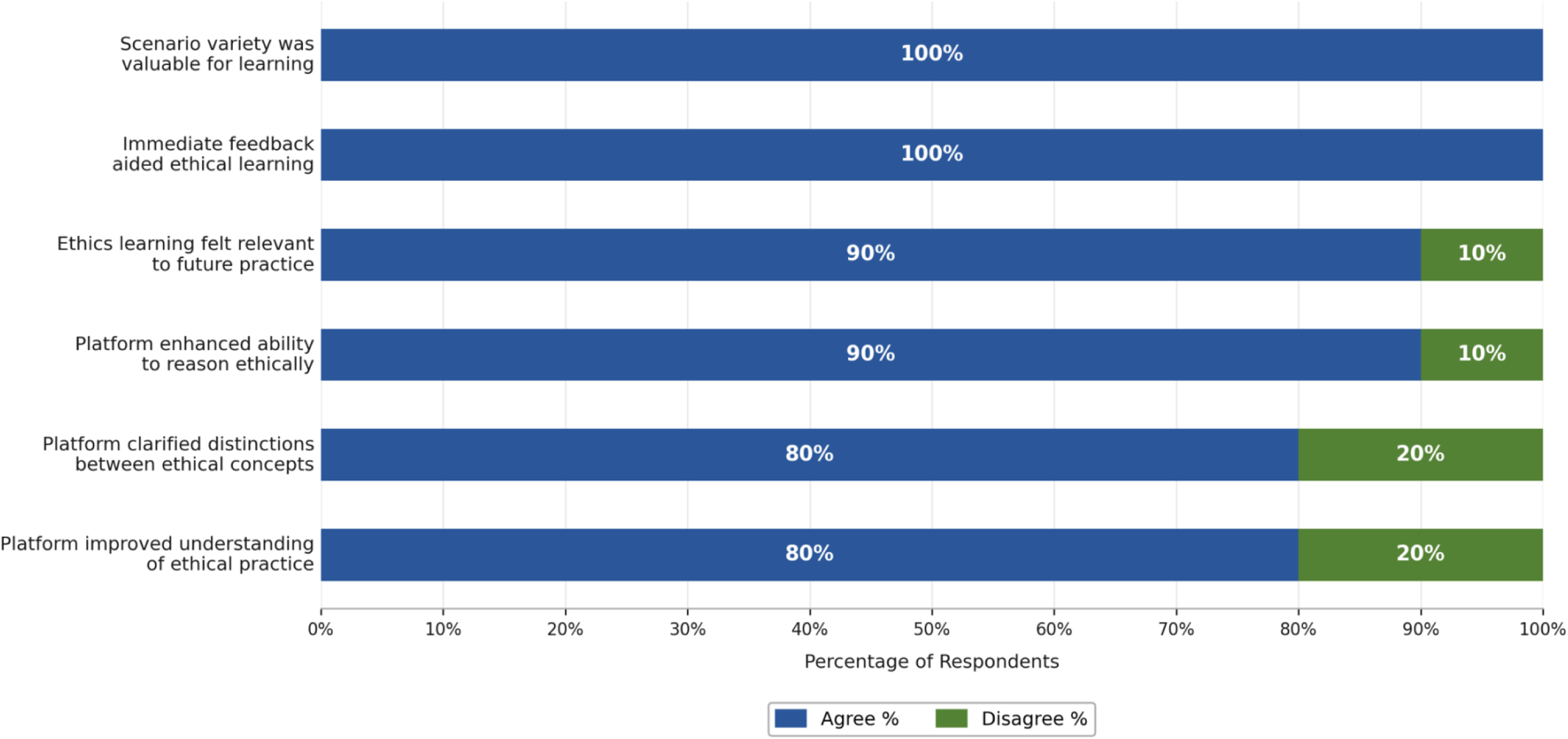
Student agreement ratings for platform utility & ethical concept learning (Objective 1)

### Objective 2: Platform usability and engagement

Five items addressed Objective 2, examining usability and the overall learning experience (Figure 3). Perceptions were again strongly positive within this small sample, with agreement from 70% to 100%. Ease of navigation reached 100% agreement; engagement with the scenarios, enjoyment of the interactive format, and willingness to reuse the platform each reached 90%. The lowest agreement (70%) concerned self-paced learning, enhancing the experience, which was the only item to receive a neutral response. Qualitatively, the most common concern was user-experience design, specifically limited control over scenario pacing and an auto-scrolling feature: nearly every response to the change-suggestion item identified pacing or auto-scrolling as problematic, with one participant noting it was difficult to read scenarios because chat boxes kept appearing and shifting the screen. Others suggested adding more questions, improving speed, and enabling navigation to review previously completed cases.

**Figure 3.**
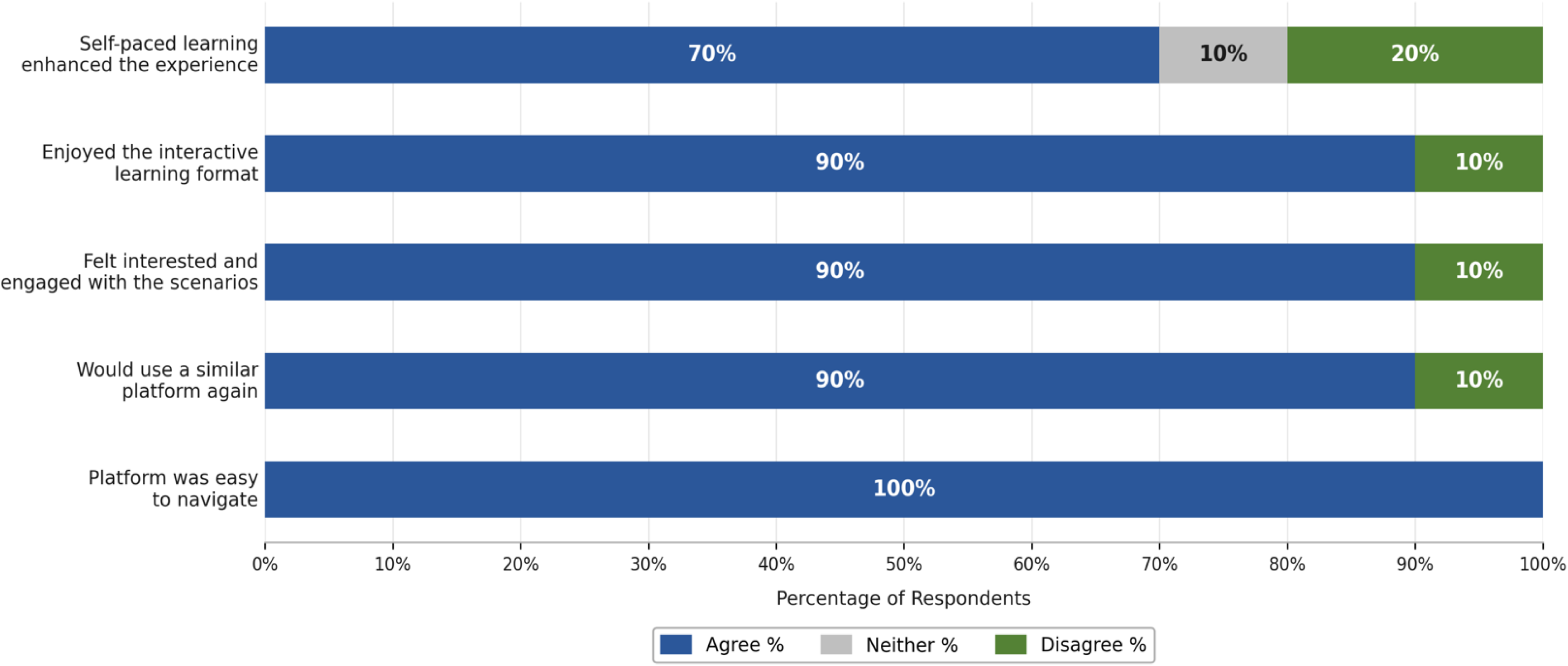
Student agreement ratings for platform usability and engagement (Objective 2)

### Objective 3: Comparison with traditional learning

Five items addressed Objective 3, comparing the platform with other learning methods, including traditional classroom instruction (Figures 4 and 5). Perceptions were more varied: agreement ranged from 45% to 90%, with higher proportions of neutral and disagreement responses than in earlier sets. The strongest agreement (90%) was for recommending this type of platform to other students; moderate agreement (67%) was for learning at one’s own pace relative to a traditional classroom. (Figure 4)

**Figure 4:**
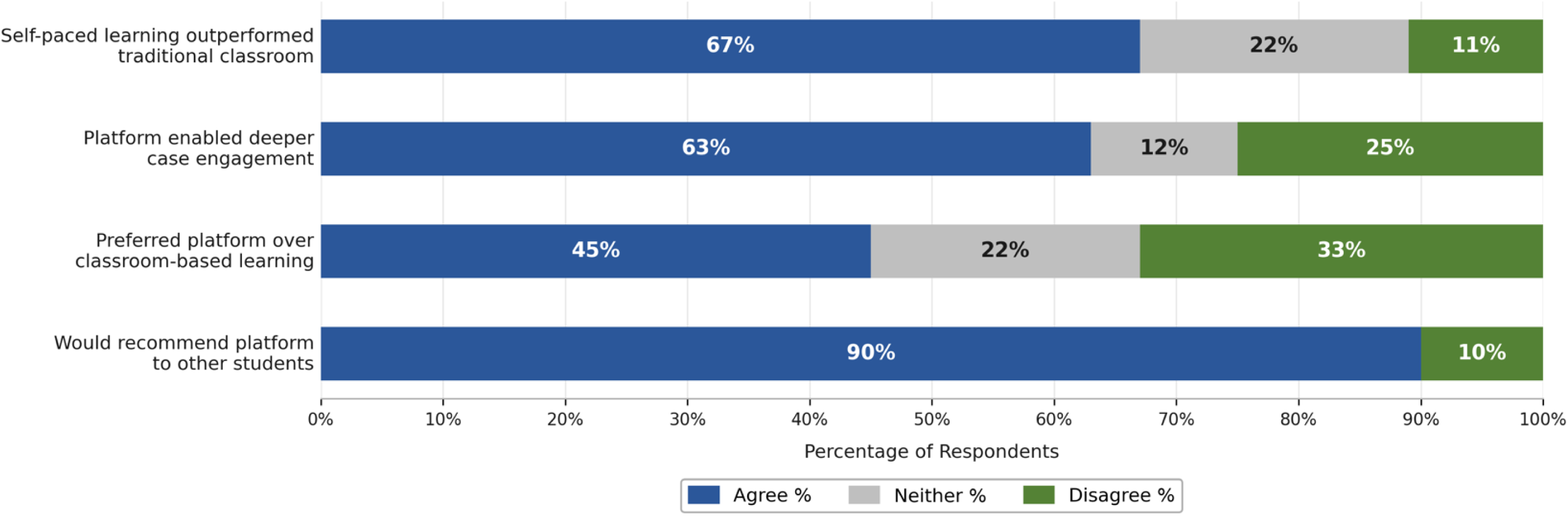
Student agreement ratings comparing Al platform to traditional learning methods. (Objective 3)

**Figure 5.**
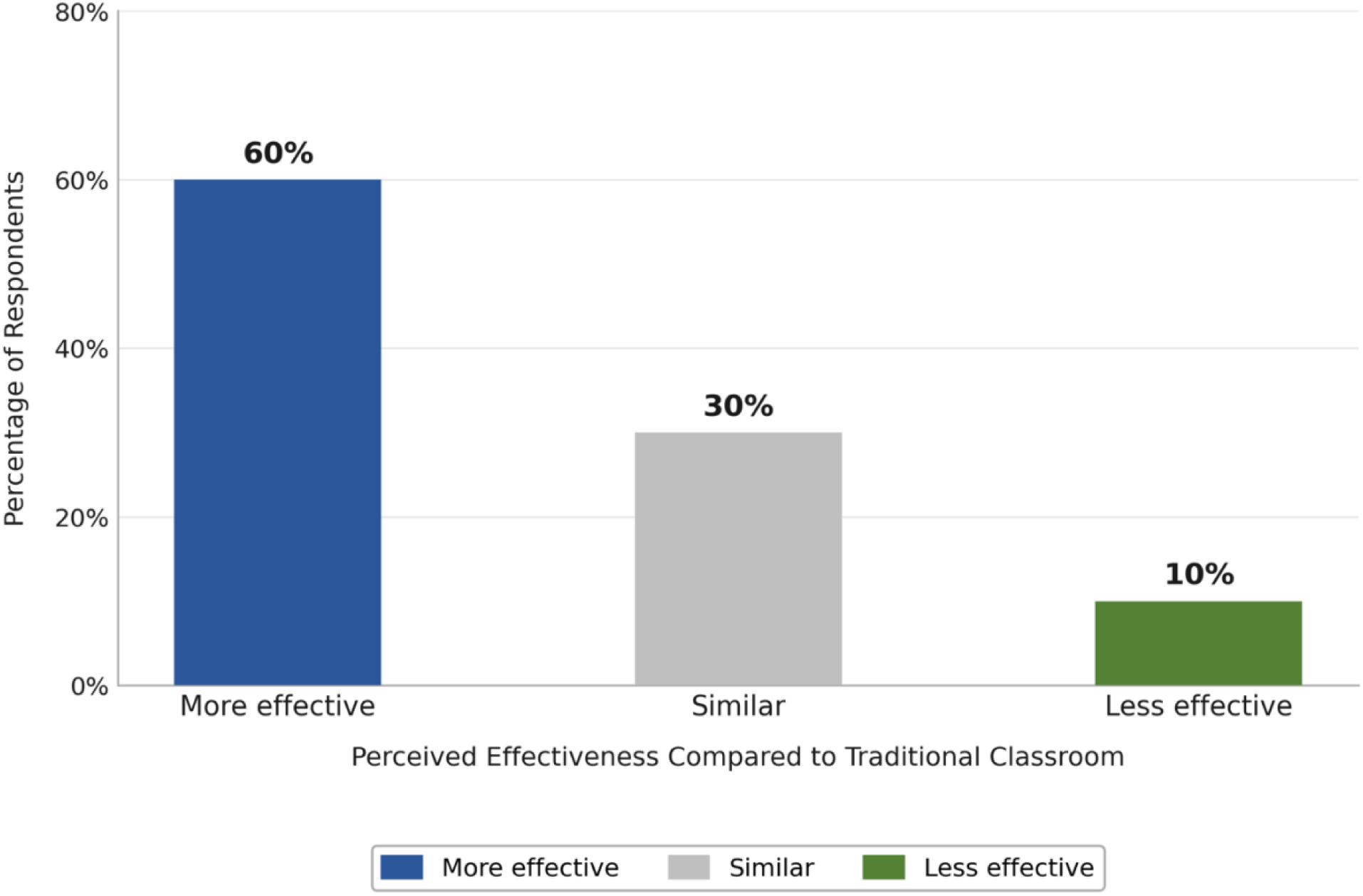
Perceived effectiveness of the AI learning platform compared with the traditional classroom method

Sixty percent (60%) of participants rated the platform as more effective than traditional learning, more than the combined total of those rating it similar or less effective (Figure 5). However, responses were divided when participants were asked whether they preferred the interactive online format to traditional case-based classroom learning, with 44% agreeing and 33% disagreeing (Figure 4). Qualitatively, most participants reported a preference for the platform, citing quick responses and live feedback; one noted ethical concepts were reinforced more effectively than in traditional settings. Others found it similar or less effective, with one characterizing it as essentially a quiz with feedback and suggesting a well-designed interactive module would be more effective.

## 5. Discussion

Within this small pilot sample, students perceived the AI-enhanced platform as a useful, engaging support for learning ethical decision-making. Across Objectives 1 and 2, participants reported high agreement on the platform’s usefulness for learning ethical concepts, engaging with scenarios, and navigating the system. All participants agreed that immediate feedback after each scenario was helpful, suggesting feedback-driven learning may play an important role in supporting understanding of ethical practice, a capability GenAI can provide at scale with instructor oversight. Strong agreement on scenario variety and the interactive format suggests such approaches may enhance engagement, consistent with reports that AI can rapidly generate realistic cases, reduce the bottleneck of expert-authored materials, and broaden the range of problems students encounter [7,9], and that AI can help students explore alternative dialogues, solutions, and perspectives within each case [8]. These observations align with active-learning approaches in which learners engage directly with realistic problems rather than passively receiving information.

At the same time, the more divided responses for Objective 3 suggest such platforms may serve best as a complement to, rather than a replacement for, traditional instruction. Although many participants reported that the platform supported engagement and self-paced learning, preferences were mixed when it was compared directly with classroom case discussions. This is consistent with arguments that AI should automate routine tasks and personalize learning so that educators can devote more time to meaningful interaction, complementing rather than replacing human-led instruction [11], and that AI can efficiently draft scenarios but that experienced tutors and clinicians must refine and review cases for realism, accuracy, and depth [9]. Students appear to continue to value real-time discussion, instructor guidance, and collaborative dialogue with peers.

Two strengths stand out for instructional design. First, the interactive, feedback-driven approach was universally valued: all participants agreed that immediate feedback supported their understanding, suggesting tailored feedback is an important design element and that a dedicated feedback-generating AI agent merits consideration in future iterations. Second, the variety of scenarios and professional contexts was rated highly, likely helping participants consider ethical issues from multiple perspectives and maintaining engagement by reducing repetition. The findings also indicate platform limitations. Agreement was somewhat lower for distinguishing the three specific ethical concepts than for general understanding, suggesting scenario-based learning may support reasoning more than it conveys theoretical distinctions, reinforcing the value of pairing it with classroom instruction. User-experience issues, principally pacing and auto-scrolling, were identified as detrimental, underscoring the need for thorough quality control and user testing of AI-generated software before deployment. Mixed views on self-paced learning further suggest that pacing control is a priority for refinement. Several study-level limitations qualify these findings. Most importantly, only 10 of an intended 175 participants responded, substantially reducing statistical reliability and generalizability. Late-term administration likely depressed participation and may have introduced response bias, as participants may have held stronger opinions or greater engagement. The absence of physiotherapy respondents means the findings should not be read as representative of all four professions. The work was also constrained by the timelines and deliverables of an undergraduate thesis project, which limited recruitment, iterative testing, and longitudinal follow-up and required discontinuing a planned refactoring of the platform into a reusable version, restricting evaluation of scalability. Finally, the study relied on self-reported perceptions rather than objective measures of learning, so results reflect attitudes toward the platform rather than confirmed gains in ethical reasoning.

Future research should examine whether such platforms produce measurable improvements in ethical reasoning beyond self-reported perceptions, ideally with larger, more diverse samples across settings and disciplines. Comparative designs contrasting fully online AI-enhanced case-based learning with hybrid models that combine interactive scenarios and classroom discussion could clarify how best to balance independent and collaborative learning, and studies of specific design features, feedback timing, scenario complexity, and opportunities for peer discussion could identify the elements that most strongly drive engagement and learning, with particular attention to user experience [8].

## 6. Conclusions

This pilot examined student perceptions of an AI-enhanced, case-based learning platform designed to support ethical reasoning among interprofessional healthcare students. Within a small cohort, students viewed the platform positively, highlighting immediate feedback, scenario variety, and interactive engagement as supports for understanding ethical concepts. The study offers an early proof of concept for applying instructor-vetted, AI-generated scenarios to interprofessional ethics education in rehabilitation-focused disciplines, where such approaches have received little prior attention. The preliminary findings also indicate that AI-enhanced platforms are best used to complement, rather than replace, traditional instruction, particularly given the usability concerns and mixed preferences for in-person interaction observed here. AI-driven case-based learning platforms hold promise for enhancing ethics education, provided future applications address the identified usability challenges and explore how best to integrate them alongside traditional approaches.

## Declaration of generative AI and AI-assisted technologies in the manuscript preparation process

During the preparation of this work the authors used Claude (Anthropic) in order to proofread, improve grammar, refine sentence structure, and enhance clarity. After using this tool, the authors reviewed and edited the content as needed and take full responsibility for the content of the published article.

## Author contributions

**Liam Rankine:** Software, Investigation, Data curation, Formal analysis, Writing – original draft, Visualization.

**Jennifer Van Bussel:** Methodology, Writing – review and editing

**Sheila Theresa Moodie:** Methodology, Writing – review and editing.

**Andrews Tawiah:** Conceptualization, Methodology, Supervision, Project administration, Writing – review and editing.

## Acknowledgements

The authors thank Dr. Ken Meadows for supporting the data collection phase of this study.

## Data availability

The data that support the findings of this study are available from the corresponding author upon reasonable request. The data are not publicly available because they contain information that could compromise the privacy of research participants.

## Code availability

The platform source code is publicly available at https://github.com/liamrankine22/Ethical-Learning-AI-Based-Case-Studies.

